# Risk of new-onset obstructive sleep apnea up to 4.5 years after COVID-19 in the urban population

**DOI:** 10.64898/2026.02.12.26346136

**Authors:** Sagar Changela, Rachel Katz, Jainam Shah, Sonya S Henry, Tim Q Duong

## Abstract

**Rationale:** Obstructive sleep apnea (OSA) is linked to cardiovascular, metabolic, and cognitive morbidity. Although COVID-19 has been associated with long-term respiratory and neurological sequelae, its role in precipitating new-onset OSA remains unclear.

**Objectives:** To evaluate whether SARS-CoV-2 infection increases risk of developing OSA up to 4.5 years post-infection and how risk varies by hospitalization status, demographics, comorbidities, and vaccination status.

**Methods:** This retrospective cohort study used electronic health records from the Montefiore Health System in the Bronx. Adults tested for SARS-CoV-2 between March 1, 2020, and August 17, 2024, were classified as hospitalized COVID+, non-hospitalized COVID+, or COVID−. Patients with prior OSA or inadequate follow-up were excluded. Inverse probability weighting adjusted for demographic, clinical, socioeconomic, and vaccination covariates. New-onset OSA was assessed using weighted Cox proportional hazards models. Secondary outcomes including hypertension, myocardial infarction, heart failure, stroke, arrhythmia, pulmonary hypertension, type 2 diabetes, and obesity were evaluated with Poisson regression. Sensitivity analysis used a pre-pandemic control cohort.

**Results:** Among 910,393 eligible patients, hospitalized [HR 1.41 (95% CI 1.14-1.73)] and non-hospitalized [HR 1.33 (95% CI 1.22-1.46)] COVID+ patients had higher adjusted risk of new-onset OSA versus COVID− controls. Similar findings were observed using historical controls (n=621046). After OSA onset, hospitalized COVID+ patients had higher risks of heart failure and pulmonary hypertension, while non-hospitalized COVID+ patients had higher risk of obesity vs COVID− patients.

**Conclusions:** SARS-CoV-2 infection is independently associated with increased risk of new-onset OSA. These findings support targeted screening in post-COVID populations.

## INTRODUCTION

Obstructive sleep apnea (OSA) is a common and serious sleep disorder characterized by repetitive episodes of upper airway collapse during sleep, leading to intermittent hypoxia, fragmented sleep, and sympathetic overactivation. It affects an estimated 10–30% of adults globally, with higher prevalence in individuals with obesity, cardiovascular disease, and metabolic disorders (1). Untreated OSA has been linked to increased risks of hypertension, stroke, type 2 diabetes, arrhythmias, and cognitive impairment (2). The pathophysiology of OSA is multifactorial and includes both anatomical factors (e.g., pharyngeal collapsibility, obesity-related airway narrowing) and non-anatomical contributors such as impaired neuromuscular control of the upper airway, altered ventilatory control, and systemic inflammation. Emerging evidence suggests that SARS-CoV-2 infection—particularly in moderate to severe cases—may unmask these vulnerabilities, thereby contributing to the development of new-onset OSA.

COVID-19 has been associated with persistent systemic inflammation, autonomic dysregulation, and central nervous system involvement (3-6), all of which can impact sleep and breathing regulation, and specifically OSA (7, 8). Furthermore, hospitalization due to COVID-19 often entails prolonged immobilization, corticosteroid use, and weight gain, which are known risk factors for OSA. Even among non-hospitalized individuals, post-acute sequelae of COVID-19 (“long COVID”) may impair respiratory function and sleep architecture, potentially precipitating the emergence of OSA (9, 10). It is unknown whether SARS-CoV-2 infection independently increases the risk of developing OSA, and who may be more susceptible. Prior studies focused on pre-existing OSA as a risk factor for acute severe COVID-19 outcomes (8), but not on COVID-19 as a potential risk for new incident OSA. Additionally, disparities in OSA diagnosis and treatment—particularly among women, younger adults, and racial and ethnic minorities—raise concerns about unequal burdens of post-COVID complications in these vulnerable groups (11-13). The role of disease severity, hospitalization status, and vaccination in modulating the risk of OSA post-infection is also unclear. While COVID-19 vaccines have demonstrated efficacy in reducing acute disease severity, their influence on long-term outcomes such as new-onset sleep disorders has not been well established, with studies focusing on effects on sleep disturbances rather than sleep disorders (14).

In light of these knowledge gaps, the present study investigated the association between SARS-CoV-2 infection and the risk of new-onset OSA using a large urban cohort up to 4.5 years post infection. The study further stratified risk by hospitalization status, demographic factors, comorbidities, and vaccination status, and validated findings using a historical comparator cohort. Understanding these relationships is essential to inform clinical screening strategies, guide post-COVID care, and improve long-term outcomes for at-risk populations.

## METHODS

### Patient sources

This retrospective cohort study was approved by the Einstein-Montefiore Institutional Review Board (2021-13658), which granted a waiver of obtaining informed consent. All methods were performed in accordance with the relevant guidelines and regulations. Clinical data were obtained from the Montefiore Health System’s electronic health records (EHR), encompassing care delivered across multiple hospitals and outpatient facilities located in the Bronx and surrounding communities. Data retrieval procedures have been described previously, and different analyses have been reported using previous versions of this large dataset (15-23).

Patient with SARS-CoV-2 PCR tests were identified between March 1, 2020, and August 17, 2024 and stratified into three groups: i) individuals who tested positive and were hospitalized for COVID-19, ii) those who tested positive but were not hospitalized, and iii) individuals who never tested positive throughout the study period. The date of the first positive result was designated as the index date for the COVID-19 positive (COVID+) cohort. Whereas the first visit after March 2020 was considered as an index date for the COVID-19 negative (COVID−) cohort. Patients were excluded if they had a documented diagnosis of OSA prior to the index date. Patients were also excluded if they did not have a follow-up visit to our health system 30 days post-index date.

### Variables

Demographic characteristics included age at index date, sex, race, and ethnicity. Socioeconomic status included insurance type and the median household income linked to the patient’s residential ZIP code. Prevalent pre-existing health conditions at index date included type-2 diabetes mellitus (T2DM), hypertension (HTN), chronic obstructive pulmonary disease (COPD), chronic kidney disease (CKD), cardiovascular disease (CVD)—defined as prior myocardial infarction, coronary artery disease, or heart failure—as well as asthma, obesity, and hypothyroidism. Vaccination status was extracted as a binary variable, defined as receipt of at least one dose of a COVID-19 vaccine prior to or during the follow-up period. This information was obtained from the Montefiore Health System immunization records and cross-referenced with New York State vaccine registries. COVID+ patients were also analyzed with respect to whether they were hospitalized during acute SARS-CoV-2 infection.

### Outcomes

Primary outcome was new incident OSA, defined by ICD-10 codes (**Supplementary Table 1)** post index date. Patients were followed until the earliest of the following: OSA diagnosis, date of death, or last recorded clinical encounter, through August 17, 2024. Secondary outcomes were new incident medical conditions post OSA diagnosis, and they included hypertension, myocardial infarction, heart failure, stroke, arrhythmia, pulmonary hypertension, type 2 diabetes, and obesity as defined ICD-10 codes. Outcomes were provided for three groups: hospitalized COVID+, non-hospitalized COVID+, and COVID−.

### Sensitivity Analysis Using Historical Cohort

As a sensitivity analysis, outcomes were analyzed using a historical cohort as controls to avoid potential misclassification in the COVID-19 negative group (e.g., undiagnosed or asymptomatic SARS-CoV-2 infection). This historical cohort comprised patients from the Montefiore Health System with index dates between January 1, 2016, and December 31, 2019— prior to the onset of the COVID-19 pandemic. Individuals with OSA diagnosed before the index date were excluded. The historical cohort was weighted using the same inverse probability weighting method described for the main analysis. Baseline characteristics were compared before and after weighting to assess covariate balance. The longest follow-up time for the historical cohort was 44 months, shorter than the 54.8-month longest follow-up of the primary cohort. As such, direct comparison of full-length survival outcomes was not appropriate. Instead, truncated hazard ratios were estimated at successive intervals (12, 24, 36, and 44 months) within the primary cohort to enable fair comparison at the 44-month time point. Cox proportional hazards models were used to estimate hazard ratios for new incident OSA in hospitalized and non-hospitalized COVID+ patients versus the historical cohort.

### Statistical Analysis

Inverse probability weighting (IPW) was applied to adjust for baseline differences across the three study groups. A multinomial logistic regression model was used to estimate the probability of each participant belonging to their observed group (hospitalized COVID-19 positive, non-hospitalized COVID-19 positive, or COVID-19 negative), based on all predefined covariates. These included age, sex, race, ethnicity, comorbidities, socioeconomic indicators, and COVID-19 vaccination status. The resulting predicted probabilities served as propensity scores (PS) used to compute inverse probability weights. Stabilized weights were calculated by dividing the overall proportion of each group by the individual’s predicted probability, using group proportions as the stabilization factor.

Covariate balance was assessed using absolute standardized mean differences, with values less than 0.2 considered acceptable. Weighted Cox proportional hazards models were then used to estimate the association between COVID-19 exposure and incident OSA. Kaplan-Meier survival curves were generated to compare time-to-event distributions across groups. Stratified Cox proportional hazard models were used to compare outcomes for different demographic subgroups. All analyses were conducted using R (version 4.4.0) and Python (version 3.8.19), with statistical significance defined as p < 0.05.

For the secondary outcomes, the risk of new-onset conditions was compared across the three exposure groups. Risk ratios and 95% confidence intervals were calculated from Poisson regression models adjusted for age, sex, race, and ethnicity.

## RESULTS

From March 2020 to August 2024, 1,389,647 unique patients visited the Montefiore Health System (**Figure 1**). There were 65,009 COVID positive (COVID+) and 1,021,351 COVID negative (COVID−) patients without past medical history of OSA, of which 57,206 COVID+ and 853,187 COVID– patients returned to our health system 30 or more days after the index date.

**Figure 1:**
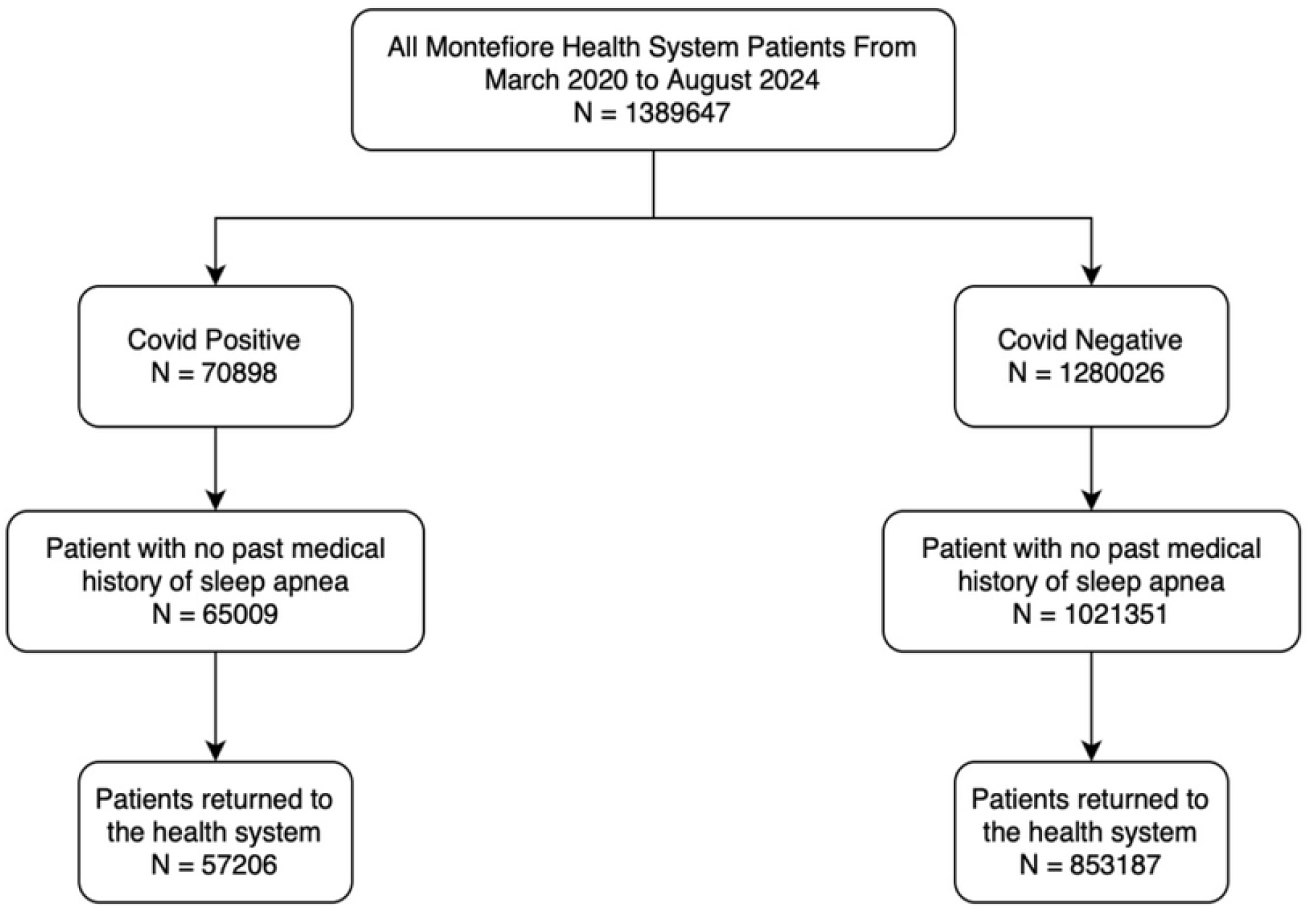
Flowchart of patient selection.

The baseline characteristics of patients with and without COVID-19 had some minor differences (**Table 1A**). COVID+ hospitalized patients were on average older (64.07 vs. 39.62 vs. 42.43 years old), were more likely to be smokers (36.8% vs 19.6% vs 19.8%), be on Medicare (35.6% vs. 7.9% vs. 11.8%) and more likely to have all major pre-existing comorbidities compared to COVID+ non-hospitalized and COVID− patients. COVID+ non-hospitalized patients were more likely to be vaccinated with at least one dose as compared to both hospitalized COVID+ and COVID− patients. Non-hospitalized COVID+ patients were more likely to have hypertension, type-2 diabetes, asthma and obesity compared to COVID− patients. To control for potential confounds, a matched cohort was created using IPW (**Table 1B**).

**Table 1:** Baseline characteristics of patients (A) prior to and (B) after inverse-probability weighting, excluding those with a history of obstructive sleep apnea. Absolute standardized differences (ASDs) were calculated to assess covariate balance across the groups. A vs C – Hospitalized COVID-19 positive vs COVID-19 negative. B vs C - Non-hospitalized COVID-19 positive vs COVID-19 negative. A vs B - Hospitalized COVID-19 positive vs Non-hospitalized COVID-19 positive

| (A) | Hospitalized<br>Covid-19<br>Positive<br>N= 15572 | Non-Hospitalized<br>Covid-19<br>Positive<br>N= 41634 | Covid-19 Negative<br>N = 853187 | Absolute Standardized Difference |  |  |
| --- | --- | --- | --- | --- | --- | --- |
|  |  |  |  | A vs C* | B vs C* | A vs B* |
| **Age, mean ± standard** | 64.07 (22.04) | 39.62 (23.54) | 42.43 (24.52) | 0.93 | 0.12 | 1.07 |
| **Age categorical) ≥ 60 years** | 10138 (65.1) | 9645 (23.2) | 244970 (28.7) | 0.78 | 0.13 | 0.93 |
| **Male, n (%)** | 7206 (46.3) | 16308 (39.2) | 375388 (44.0) | 0.05 | 0.10 | 0.14 |
| **Race, n (%)** |  |  |  |  |  |  |
| White | 2046 (13.1) | 4287 (10.3) | 115744 (13.6) | 0.01 | 0.10 | 0.09 |
| Black | 5403 (34.7) | 12774 (30.7) | 227635 (26.7) | 0.17 | 0.10 | 0.09 |
| Asian | 556 (3.6) | 2111 (5.1) | 26539 (3.1) | 0.03 | 0.10 | 0.07 |
| Others | 7567 (48.6) | 22462 (54.0) | 483269 (56.6) | 0.16 | 0.05 | 0.11 |
| **Ethnicity Hispanic, n (%)** | 7363 (47.3) | 22314 (53.6) | 484569 (56.8) | 0.19 | 0.06 | 0.13 |
| **ZIP Code Median Income, n** |  |  |  |  |  |  |
| 1st Quartile (0-25) | 3826 (24.6) | 11728 (28.2) | 280818 (32.9) | 0.18 | 0.10 | 0.08 |
| 2nd Quartile (25-50) | 4499 (28.9) | 10826 (26.0) | 193760 (22.7) | 0.14 | 0.07 | 0.06 |
| 3rd Quartile (50 -75) | 4678 (30.0) | 11198 (26.9) | 199962 (23.4) | 0.15 | 0.08 | 0.07 |
| 4th Quartile (75-100) | 2569 (16.5) | 7882 (18.9) | 178647 (20.9) | 0.11 | 0.05 | 0.06 |
| **Insurance, n (%)** |  |  |  |  |  |  |
| Medicare | 5511 (35.6) | 3278 (7.9) | 96762 (11.8) | 0.58 | 0.13 | 0.71 |
| Medicaid | 5525 (35.7) | 19038 (46.1) | 366808 (44.7) | 0.18 | 0.03 | 0.21 |
| Private | 3870 (25.0) | 14183 (34.3) | 266431 (32.5) | 0.16 | 0.04 | 0.21 |
| Self-pay | 572 (3.7) | 4840 (11.7) | 90861 (11.1) | 0.28 | 0.02 | 0.30 |
| **Smoking** | 5727 (36.8) | 8172 (19.6) | 168936 (19.8) | 0.38 | 0.004 | 0.39 |
| **Vaccination (at least one dose)** | 6494 (41.7) | 21485 (51.6) | 304486 (35.7) | 0.12 | 0.33 | 0.2 |
| **Comorbidities, n (%)** |  |  |  |  |  |  |
| Hypertension | 10775 (69.2) | 11398 (27.4) | 172252 (20.2) | 1.13 | 0.17 | 0.92 |
| Type-2 Diabetes | 8072 (51.8) | 8768 (21.1) | 115663 (13.6) | 0.89 | 0.20 | 0.68 |
| Cardiovascular disease | 5616 (36.1) | 3005 (7.2) | 45568 (5.3) | 0.82 | 0.08 | 0.75 |
| COPD | 1912 (12.3) | 887 (2.1) | 11003 (1.3) | 0.45 | 0.07 | 0.40 |
| Asthma | 3126 (20.1) | 7574 (18.2) | 77587 (9.1) | 0.32 | 0.27 | 0.05 |
| Obesity | 4613 (29.6) | 7894 (19.0) | 91894 (10.8) | 0.48 | 0.23 | 0.25 |
| Chronic Kidney Disease | 5078 (32.6) | 2573 (6.2) | 34440 (4.0) | 0.80 | 0.09 | 0.71 |
| Hypothyroidism | 1685 (10.8) | 1844 (4.4) | 23189 (2.7) | 0.33 | 0.09 | 0.24 |
| **Outcome** |  |  |  |  |  |  |
| Obstructive sleep apnea | 327 (2.09%) | 726 (1.74%) | 12046 (1.41%) | - | - | - |

| (B) | Hospitalized<br>Covid-19 | Non-Hospitalized | Covid-19 Negative<br>N = 853459 | Absolute Standardized Difference |  |  |
| --- | --- | --- | --- | --- | --- | --- |
|  |  |  |  | A vs C* | B vs C* | A vs B* |
| **Age, mean ± standard** | 43.72 (24.89) | 42.16 (23.72) | 42.69 (24.65) | 0.04 | 0.02 | 0.06 |
| **Age categorical) ≥ 60 years** | 4051 (29.2) | 10777 (26.1) | 249534 (29.2) | 0.001 | 0.07 | 0.07 |
| **Male, n (%)** | 5569 (40.2) | 18323 (44.3) | 373908 (43.8) | 0.07 | 0.01 | 0.08 |
| **Race, n (%)** |  |  |  |  |  |  |
| White | 1829 (13.2) | 5448 (13.2) | 114446 (13.4) | 0.006 | 0.007 | 0.001 |
| Black | 3864 (27.9) | 11323 (27.4) | 230520 (27.0) | 0.02 | 0.009 | 0.01 |
| Asian | 435 (3.1) | 1363 (3.3) | 27387 (3.2) | 0.004 | 0.005 | 0.009 |
| Others | 7732 (55.8) | 23181 (56.1) | 481109 (56.4) | 0.01 | 0.005 | 0.006 |
| **Ethnicity Hispanic, n (%)** | 7881 (56.9) | 23149 (56.0) | 481993 (56.5) | 0.008 | 0.009 | 0.02 |
| **ZIP Code Median Income, n** |  |  |  |  |  |  |
| 1st Quartile (0-25) | 4759 (34.3) | 13341 (32.3) | 277802 (32.6) | 0.04 | 0.006 | 0.04 |
| 2nd Quartile (25-50) | 3166 (22.8) | 9624 (23.3) | 196052 (23.0) | 0.003 | 0.008 | 0.01 |
| 3rd Quartile (50 -75) | 3293 (23.8) | 9769 (23.6) | 202383 (23.7) | 0.001 | 0.002 | 0.03 |
| 4th Quartile (75-100) | 2642 (19.1) | 8582 (20.8) | 177225 (20.8) | 0.04 | 0.001 | 0.04 |
| **Insurance, n (%)** |  |  |  |  |  |  |
| Medicare | 1780 (12.8) | 4530 (11.0) | 99121 (11.6) | 0.04 | 0.02 | 0.06 |
| Medicaid | 5858 (42.3) | 17856 (43.2) | 366805 (43.0) | 0.01 | 0.005 | 0.02 |
| Private | 4491 (32.4) | 13153 (31.8) | 266672 (31.2) | 0.03 | 0.01 | 0.01 |
| Self-pay | 1731 (12.5) | 5777 (14.0) | 120864 (14.2) | 0.05 | 0.005 | 0.04 |
| **Smoking** | 3292 (23.8) | 8119 (19.7) | 171525 (20.1) | 0.09 | 0.01 | 0.1 |
| **Vaccination (at least one dose)** | 4896 (35.3) | 15421 (37.5) | 311668 (36.5) | 0.03 | 0.02 | 0.05 |
| **Comorbidities, n (%)** |  |  |  |  |  |  |
| Hypertension | 3569 (25.8) | 8962 (21.7) | 182517 (21.4) | 0.1 | 0.007 | 0.09 |
| Type-2 Diabetes | 2685 (19.4) | 6163 (14.9) | 124499 (14.6) | 0.13 | 0.009 | 0.12 |
| Cardiovascular disease | 1075 (7.8) | 2429 (5.9) | 51029 (6.0) | 0.07 | 0.004 | 0.07 |
| COPD | 339 (2.4) | 652 (1.6) | 13135 (1.5) | 0.06 | 0.003 | 0.06 |
| Asthma | 1986 (14.3) | 4114 (10.0) | 83002 (9.7) | 0.14 | 0.008 | 0.13 |
| Obesity | 2431 (17.5) | 4864 (11.8) | 98111 (11.5) | 0.17 | 0.009 | 0.16 |
| Chronic Kidney Disease | 863 (6.2) | 1965 (4.8) | 39709 (4.7) | 0.07 | 0.005 | 0.06 |
| Hypothyroidism | 629 (4.5) | 1251 (3.0) | 25176 (2.9) | 0.08 | 0.005 | 0.08 |

Figure 2. presents the Kaplan-Meier analysis for new-onset OSA before and after IPW. In the unmatched cohort, hospitalized and non-hospitalized COVID+ patients had higher cumulative incidences of new-onset OSA compared to COVID− patients (contemporary controls). However, the cumulative incidences were similar between hospitalized and non-hospitalized COVID+ cohorts. With IPW, the differences in cumulative incidences between hospitalized and non-hospitalized COVID+ patients to COVID− patients became smaller. With IPW, the COVID+ hospitalized patients [Cox-proportional HR 1.41 (95% CI 1.14-1.73)] and COVID+ non-hospitalized patients [HR 1.33 (95% CI 1.22-1.46)] were at the higher risk of OSA as compared to COVID− patients.

**Figure 2:**
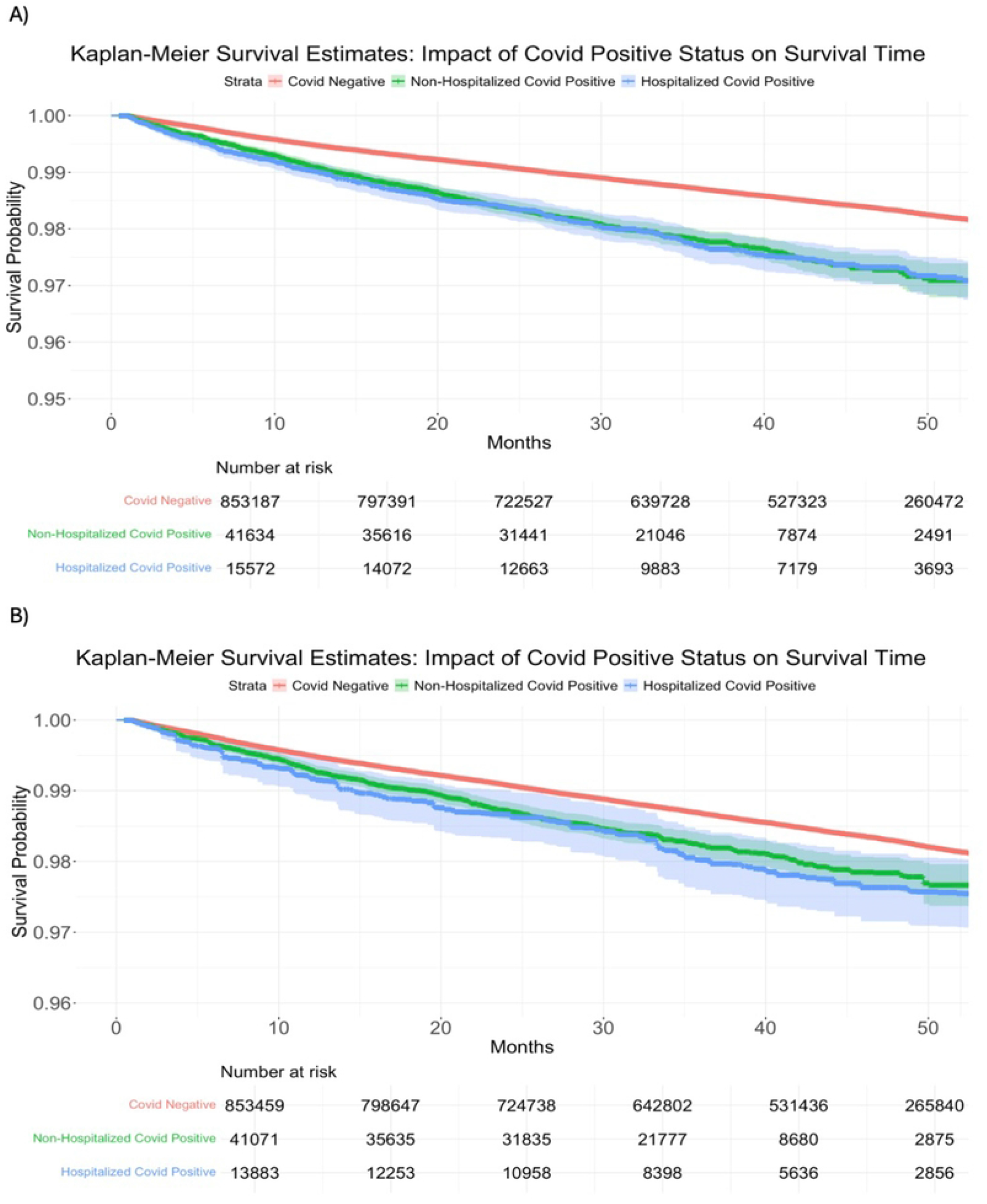
Kaplan-Meier survival curve for outcome obstructive sleep apnea. A) Unmatched data B) Inverse probability weighted data.

**Table 2** shows the new-onset secondary outcomes after the OSA diagnosis. Hospitalized COVID+ patients had higher new incidence of heart failure (8.42%), stroke (3.68%), and arrythmia (8.16%), pulmonary hypertension (5.08%) and type 2 diabetes (12.72%) than non-hospitalized COVID+ patients and COVID− patients (contemporary controls). Non-hospitalized COVID+ patients had higher new incidence of obesity (32.40%) compared to hospitalized COVID+ patients and COVID− patients. COVID− patients had a higher onset of hypertension (9.69%) compared to hospitalized and non-hospitalized COVID+ patients. The hospitalized COVID+ positive patients had a statistically significant higher adjusted risk of developing heart failure [2.33 CI (1.46-3.73)] and pulmonary hypertension [1.98 CI (1.16-3.39)] than COVID− patients. Non-hospitalized COVID+ patients had a statistically significant higher risk of developing obesity [1.16 CI (1.03-1.32)] than COVID− patients.

**Table 2:**
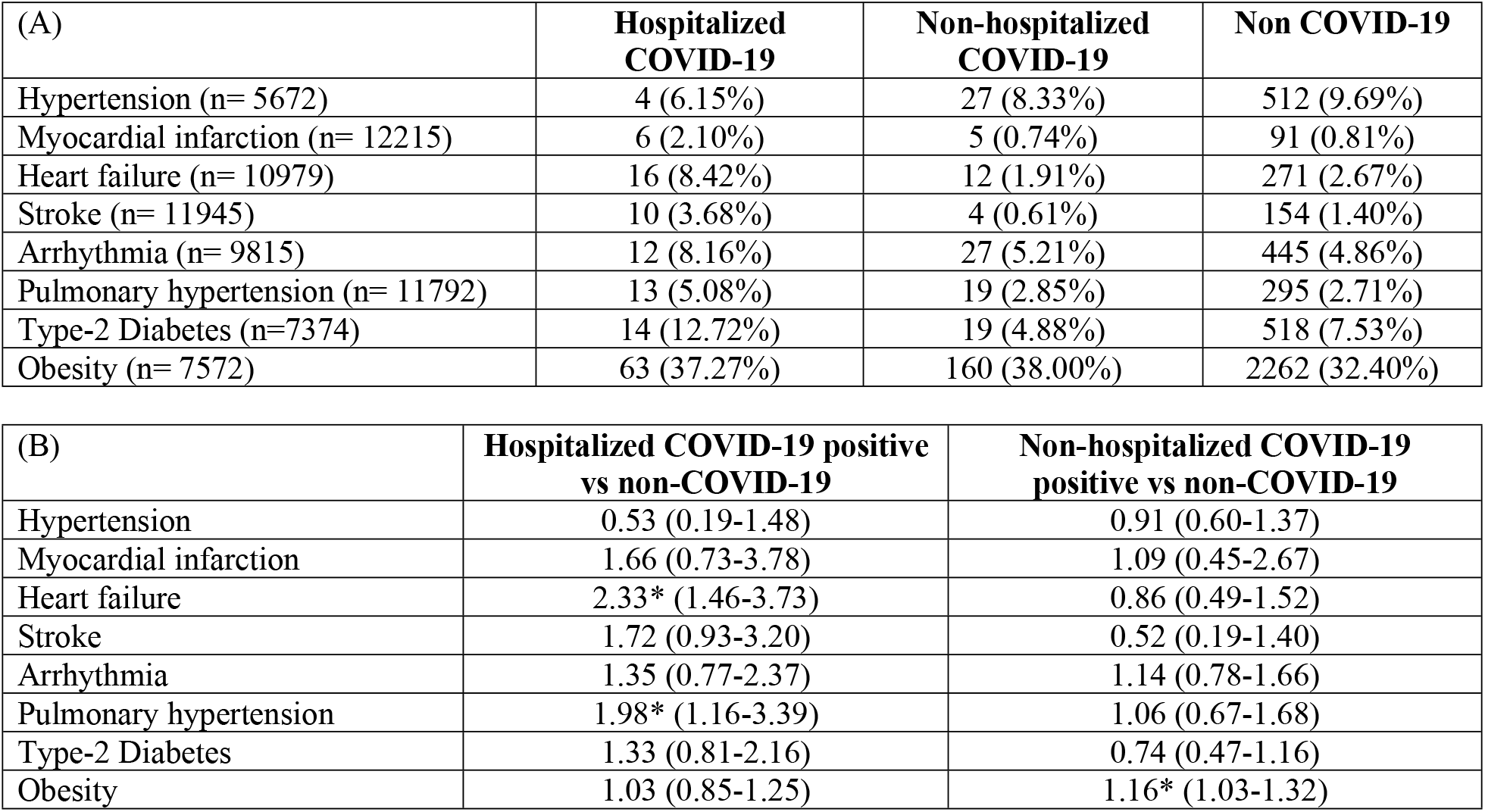
(A) New-Onset Secondary Health Conditions Post Obstructive Sleep Apnea, and (B) Risk ratio of after with adjustment for age, gender, race, and ethnicity.

| (A) | **Hospitalized COVID-19** | **Non-hospitalized COVID-19** | **Non COVID-19** |
| --- | --- | --- | --- |
| Hypertension (n= 5672) | 4 (6.15%) | 27 (8.33%) | 512 (9.69%) |
| Myocardial infarction (n= 12215) | 6 (2.10%) | 5 (0.74%) | 91 (0.81%) |
| Heart failure (n= 10979) | 16 (8.42%) | 12 (1.91%) | 271 (2.67%) |
| Stroke (n= 11945) | 10 (3.68%) | 4 (0.61%) | 154 (1.40%) |
| Arrhythmia (n= 9815) | 12 (8.16%) | 27 (5.21%) | 445 (4.86%) |
| Pulmonary hypertension (n= 11792) | 13 (5.08%) | 19 (2.85%) | 295 (2.71%) |
| Type-2 Diabetes (n=7374) | 14 (12.72%) | 19 (4.88%) | 518 (7.53%) |
| Obesity (n= 7572) | 63 (37.27%) | 160 (38.00%) | 2262 (32.40%) |

| (B) | **Hospitalized COVID-19 positive vs non-COVID-19** | **Non-hospitalized COVID-19 positive vs non-COVID-19** |
| --- | --- | --- |
| Hypertension | 0.53 (0.19-1.48) | 0.91 (0.60-1.37) |
| Myocardial infarction | 1.66 (0.73-3.78) | 1.09 (0.45-2.67) |
| Heart failure | 2.33* (1.46-3.73) | 0.86 (0.49-1.52) |
| Stroke | 1.72 (0.93-3.20) | 0.52 (0.19-1.40) |
| Arrhythmia | 1.35 (0.77-2.37) | 1.14 (0.78-1.66) |
| Pulmonary hypertension | 1.98* (1.16-3.39) | 1.06 (0.67-1.68) |
| Type-2 Diabetes | 1.33 (0.81-2.16) | 0.74 (0.47-1.16) |
| Obesity | 1.03 (0.85-1.25) | 1.16* (1.03-1.32) |

Analysis for different demographic sub-groups (**Figure 3**) showed that the association between hospitalized COVID+ and new-onset OSA was more pronounced in patients younger than 60 years, Black (as compared to White), and in asthmatic patients. The association between non-hospitalized COVID+ patients and new-onset OSA was more pronounced in females (as compared to males), Hispanics (as compared to non-Hispanics), and patients with all major comorbidities. Vaccinated and unvaccinated patients showed a similar level of risk of new-onset OSA among hospitalized and non-hospitalized COVID+ patients.

**Figure 3:**
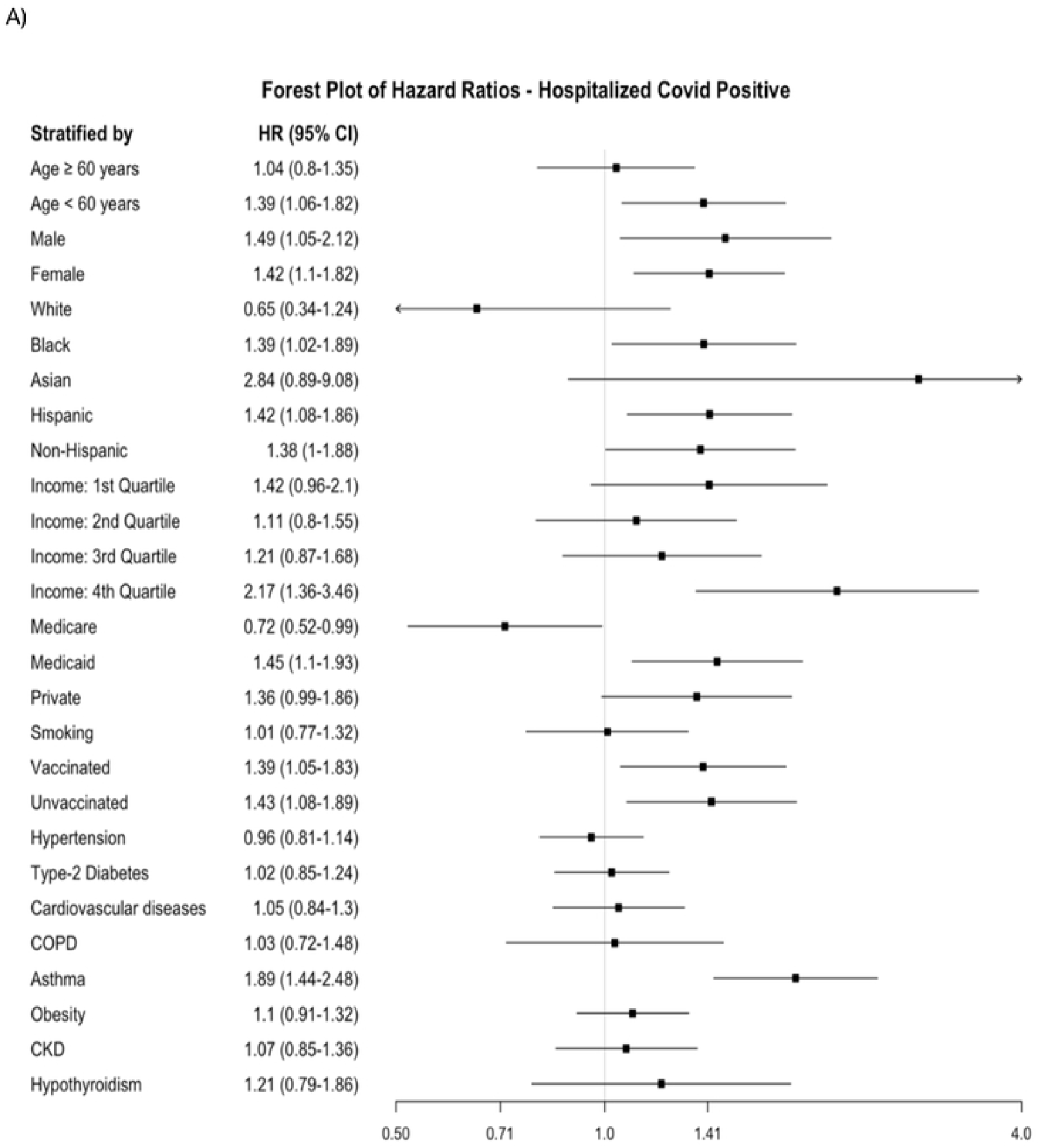
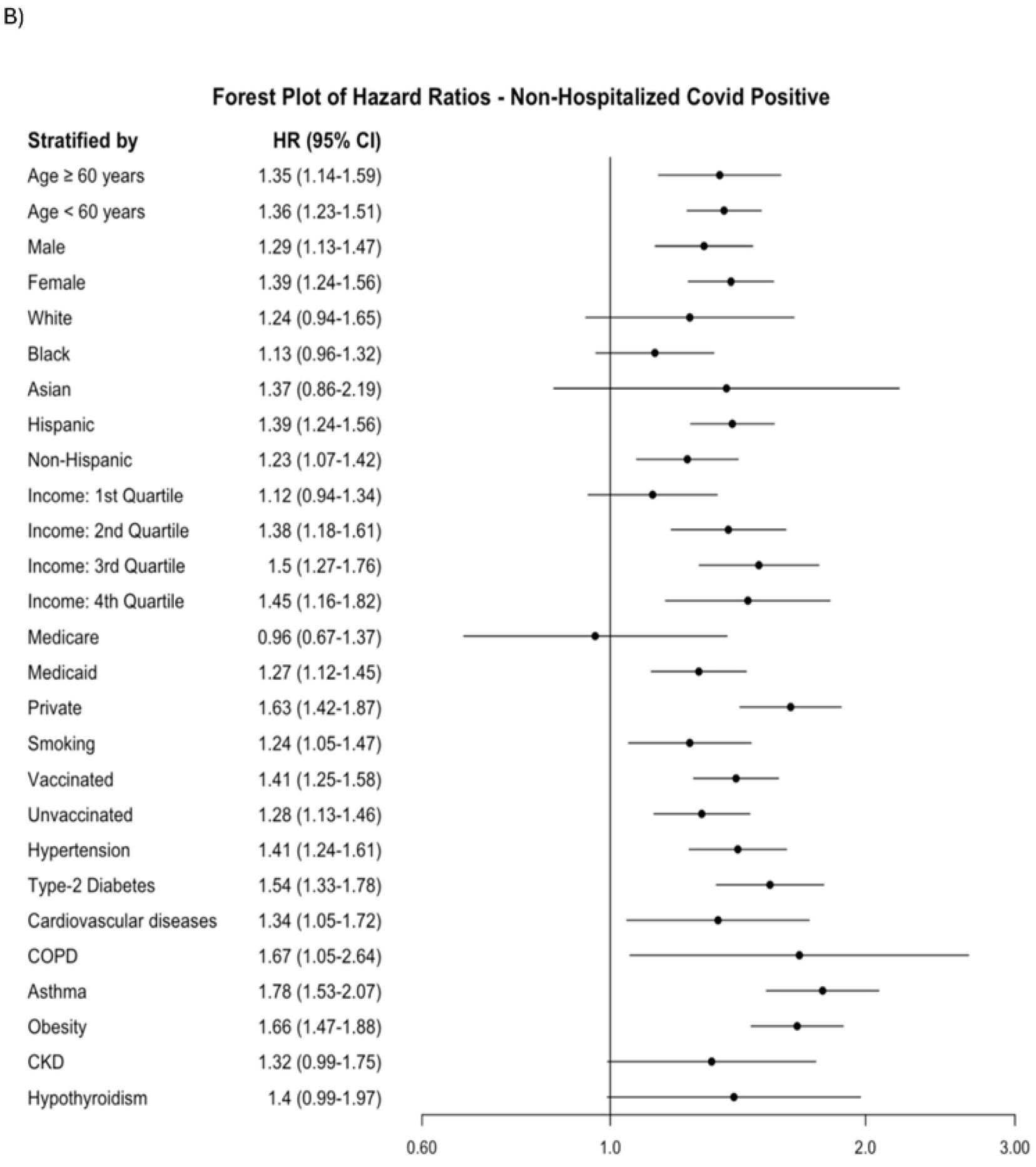
Stratified analysis for (A) Hospitalized Covid-19 Positive vs Covid Negative, and (B) Non-Hospitalized Covid-19 Positive vs Covid Negative

As a sensitivity analysis, comparisons were made with the historical cohort as control (**Appendix 1**). The COVID+ hospitalized patients [HR 2.09 (95% CI 1.73-2.55)] and COVID+ non-hospitalized patients [HR 1.56 (95% CI 1.42-1.71)] were at a higher risk of OSA compared to the historical cohort. The overall findings were consistent with the main analysis that used a contemporary cohort as control.

## DISCUSSION

This study investigated the risk of new-onset OSA associated with SARS-CoV-2 infection. Both hospitalized [HR 1.41, 95% CI: 1.14–1.73] and non-hospitalized [HR 1.33, 95% CI: 1.22–1.46] COVID+ patients demonstrated an increased risk of new-onset OSA compared to COVID− individuals. Sensitivity analysis using a historical cohort confirmed these findings. Subgroup analysis revealed stronger associations in younger patients, Black individuals, and those with asthma (hospitalized), and in females, Hispanics, and those with multiple comorbidities (non-hospitalized). COVID-19 vaccination status did not significantly affect OSA risk.

### Association Between COVID-19 and New-Onset OSA

The increased risk of OSA among both hospitalized and non-hospitalized COVID+ individuals supports the hypothesis that SARS-CoV-2 contributes to or unmasks latent vulnerabilities to sleep-disordered breathing. The association between SARS-CoV-2 infection and increased risk of new-onset OSA highlights a critical intersection between viral pathophysiology and sleep-disordered breathing. While OSA has traditionally been viewed as a chronic condition driven by anatomical and neuromuscular factors, there is evidence suggesting acute illnesses like COVID-19 can act as triggers or accelerants of underlying vulnerabilities (8, 10). One proposed mechanism is sustained low-grade systemic inflammation following SARS-CoV-2 infection, which may impair upper airway neuromuscular control and reduce pharyngeal muscle tone, predisposing individuals to airway collapse during sleep (7). Elevated levels of inflammatory cytokines such as IL-6 and TNF-α (24)—commonly observed in post-acute COVID-19 syndromes—have been shown to affect both respiratory drive and upper airway stability, mechanisms central to the pathogenesis of OSA.

Moreover, COVID-19 has been associated with significant dysregulation of the autonomic nervous system, including persistent sympathetic overactivity, which is also a known contributor to both OSA and its cardiovascular consequences (25-28). SARS-CoV-2 neurotropism may further exacerbate this risk: neuropathological studies and imaging have demonstrated the virus’s potential to impact the brainstem, including regions involved in central respiratory pattern generation (29). Disruption of these central pathways may impair ventilatory control and lead to the emergence of central or mixed apneic events, particularly in patients with predisposing comorbidities.

Additionally, changes in body composition and weight associated with COVID-19 illness may play a role. Some patients experience significant weight gain due to reduced mobility during recovery or post-infectious fatigue, which can alter the mechanical properties of the upper airway. Importantly, muscle wasting in respiratory and pharyngeal muscles as a result of severe COVID-19 illness could reduce airway patency, thereby contributing to OSA development in patients without traditional risk factors (30, 31).

From a clinical standpoint, the emergence of OSA post-COVID may not only reflect a biological consequence of the infection but also a heightened detection bias, as increased awareness of post-acute sequelae prompts more rigorous sleep evaluations. However, the persistence of increased risk even after adjusting for healthcare utilization in many studies suggests that the relationship is likely more than observational artifact. Furthermore, evidence from longitudinal cohorts suggests that sleep-related symptoms—such as non-restorative sleep, fatigue, and hypersomnia (32, 33)—are commonly reported features of long COVID, further supporting a mechanistic link between the infection and sleep-disordered breathing.

Taken together, these findings underscore the need for multidisciplinary surveillance and risk stratification in post-COVID care. Screening for OSA should be considered in patients presenting with persistent fatigue, insomnia, neurocognitive dysfunction, or cardiopulmonary symptoms, even in the absence of classic OSA risk factors such as obesity. Given the long-term consequences of untreated OSA and the elevated risk observed even in non-hospitalized individuals, targeted screening in post-COVID patients, particularly among women, racial and ethnic minorities, and those with comorbidities, may be warranted.

### Demographic and Clinical Subgroup Findings

The differential risk profiles observed across demographic and clinical subgroups underscore the importance of considering intersecting vulnerabilities in post-COVID care. The heightened risk among younger individuals and Black patients in the hospitalized cohort may reflect a combination of physiological, socioeconomic, and access-related factors. Notably, Black populations in the U.S. have historically shown higher rates of undiagnosed and untreated OSA (11), and this disparity may be exacerbated in the post-COVID context due to unequal health system engagement during the pandemic (13).

In the non-hospitalized cohort, the elevated risk among females and Hispanics also warrants close attention. Although OSA is traditionally more prevalent in males, underdiagnosis in women due to atypical symptom presentation is well-documented (34). COVID-19 may serve as a catalyst for clinical evaluation and diagnosis in these groups. Additionally, the association with multiple comorbidities supports the notion that pre-existing systemic vulnerabilities—such as cardiovascular or metabolic disorders—may amplify the risk of OSA in the aftermath of COVID-19 (35).

Vaccination status did not significantly affect the risk of new-onset OSA. While vaccines are known to reduce acute severity and mortality of COVID-19, their impact on long-term sequelae, particularly neuroinflammatory or structural changes linked to sleep disorders, is not well characterized (14). The lack of a protective effect in this domain may suggest that the mechanisms driving post-COVID OSA are not entirely mitigated by viral load reduction, pointing toward broader systemic or host-response factors.

### Limitations

Our retrospective study has several limitations. Our data were limited to those who returned to our health system for care over the study period and it is possible patients, who had more severe COVID-19 or had more urgent health concerns, were more likely to return to our health system. However, return care included any visit, including regular medical checkup in both COVID+ and COVID− cohorts. This limitation was addressed by performing sensitivity analysis by comparing patient profiles of those who returned those who did not. The patient characteristics were similar between the two groups. This study relied on the accuracy of the electronic health record, which in datasets of this size could result in inaccuracies or misdocumentations going unnoticed. To ensure accuracy, routine manual chart review of any suspicious variables on subsets of patients were performed over the last few years. Home SARS-CoV-2 tests were not used because they were not reliably documented in our electronic health record. Exclusive reliance on PCR tests in our health system could have led to patients being misclassified as COVID− because patients were tested elsewhere. Such contamination of the COVID− group with COVID-exposed patients likely underestimated any potential impact of the infection. However, cases of severe COVID-19 were unlikely to have been missed due to the need for inpatient admission, which could have only occurred at our health system, the largest in the Bronx with no other major healthcare providers. Furthermore, the sensitivity analysis using (pre-2019) historical controls found similar results when comparing to COVID−. Associations of various COVID-19 treatments with outcomes were not studied because there were many combinations of treatments which were heterogeneous and not systematically administered throughout time, especially during the early pandemic. Outcomes with respect to different waves/strains were not studied because there were mixtures of strains and strains did not necessarily reflect severity. Instead, hospitalization or IMV/IUC status was used as an indicator of COVID-19 severity. Future studies incorporating polysomnography and OSA severity indices will be essential to differentiate between mild, transient, and clinically significant post-COVID sleep-disordered breathing. This is a study from a single health system with multiple hospitals and outpatient clinics with over 7.5 million unique patient encounters per year. This health system is the predominant health system in the Bronx and its environs with no other comparable size healthcare providers. While our conclusions applied to the population in our health system, they might not be generalizable to the entire population in the Bronx and beyond. As our cohort is diverse, our findings may not be representative of less diverse populations. Future research should include multicenter studies to enhance generalizability. Retrospective analysis provides association, not cause-and-effect, between exposure and outcome. The exposure might not have led to outcome or if an undetected factor could have influenced both. Lastly, although all major confounds using multivariate analysis were corrected for, there could be unknown unintentional patient selection biases and residual confounds.

## CONCLUSIONS

This study provides compelling evidence that SARS-CoV-2 infection is associated with an increased risk of developing new-onset obstructive sleep apnea, regardless of hospitalization status. The elevated risk among younger individuals, racial and ethnic minorities, and patients with asthma or multiple comorbidities suggests that COVID-19 may exacerbate underlying vulnerabilities in sleep-disordered breathing. These findings highlight the need for heightened clinical vigilance for OSA-related complaints in patients recovering from COVID-19, especially those with known risk factors or who experienced hospitalization. Given the significant cardiovascular and cognitive risks associated with untreated OSA, early screening and intervention may be warranted. Furthermore, future research should explore the pathophysiological mechanisms linking COVID-19 to sleep-disordered breathing, potentially via neuroimaging, inflammatory marker profiling, and longitudinal follow-up.

## Data Availability

Data can be provided upon reasonable request.

## Author Contribution

Designing the Study - SC, TQD

Data collection and statistical analysis – SC

Creating tables and figures - SC

Data acquisition - SH

Interpretation of results - SC, SH, TQD

Drafting manuscript - RK, JS, SC

Editing Manuscript and Supervising - TQD

